# Addition of Tocilizumab to the standard of care reduces mortality in severe COVID-19: A systematic review and meta-analysis

**DOI:** 10.1101/2020.07.10.20150680

**Authors:** Umesha Boregowda, Abhilash Perisetti, Arpitha Nanjappa, Mahesh Gajendran, Hemant Goyal

## Abstract

**Introduction:** Tocilizumab is an anti-interleukin-6 antibody that has been used for the treatment of severe coronavirus disease 2019 (COVID-19). However, the concrete evidence of its benefit in reducing the mortality in severe COVID-19 is lacking. Therefore, we performed a systematic review and meta-analysis of relevant studies that compared the efficacy of Tocilizumab in severe COVID-19 vs. standard of care alone.

**Methods:** Literature search for studies that compared ‘Tocilizumab’ and ‘Standard of care’ in the treatment of COVID-19 was done using major online databases from December 2019 to June 14^th^, 2020. Search words ‘ Tocilizumab,’ ‘anti-interleukin-6 antibody,’ and ‘COVID-19’ or ‘coronavirus 2019’ in various combinations were used. Articles in the form of abstracts, letters without original data, case reports, and reviews were excluded. Data was gathered on an excel sheet, and statistical analysis was performed using Review Manager 5.3.

**Results:** Sixteen studies were eligible from 693 initial studies, including 3,641 patients (64% males). There were thirteen retrospective studies and three prospective studies. There were 2,488 patients in the standard of care group (61.7%) and 1,153 patients (68.7%) in the Tocilizumab group. The death rate in the tocilizumab group, 22.4% (258/1153), was lower than the standard of care group, 26.21% (652/2,488) (Pooled odds ratio 0.57 [95% CI 0.36-0.92] p=0.02). There was a significant heterogeneity (Inconsistency index= 80%) among the included studies.

**Conclusion:** The addition of Tocilizumab to the standard of care might reduce the mortality in severe COVID-19. Larger randomized clinical trials are needed to validate these findings.

## INTRODUCTION

Coronavirus Disease 2019 (COVID-19) is a viral disease caused by Severe acute respiratory syndrome coronavirus-2 (SARS-CoV-2) that originated from Wuhan city of Hubei province in China in December 2019.[1, 2] COVID-19 has spread around the world affecting more than 9 million people with more than 473,000 deaths globally.[3] About 6-10% of COVID-19 patients develop acute respiratory distress syndrome (ARDS) with a high mortality rate.[4, 5] The virus commonly spreads through droplets. However, the virus has also been found in gastrointestinal secretions.[6-8] Therefore, potentially the virus transmission could happen through the fecal-oral route as well. Common clinical symptoms of COVID-19 include fever, cough, malaise, shortness of breath, and fatigue.[9] Gastrointestinal manifestations such as diarrhea, and abnormal liver chemistries have also been observed.[10, 11] Laboratory abnormalities in COVID-19 are increased inflammatory markers such as C-reactive protein, ferritin, erythrocyte sedimentation rate, Interleukin-6 (IL-6), and lymphocytopenia, among others.[12] COVID-19 has posed tough challenges for healthcare workers because of the absence of effective and proven treatment. COVID-19 has a high case fatality rate of 4.5% among patients older than 60 years.[13] As a result, various antiviral and anti-inflammatory medications such as hydroxychloroquine, remdesivir, monoclonal antibodies, and convalescent serum are being used on a compassionate basis in patients with severe COVID-19.[14-16] Currently treatment of severe COVID-19 with antiviral therapy such as remdesivir, hydroxychloroquine, lopinavir, ritonavir, or any other antiviral agents along with supportive treatment is considered as the standard of care (SOC). Recently steroids such methylprednisolone has been included as standard of treatment.[17]

ARDS is characterized by inflammatory cytokine release syndrome, among which IL-6 has a pivotal role.[18] IL-6 is one of the significant inflammatory markers released during the cytokine storm due to COVID-19.[19]. Tocilizumab (TCZ) has been proposed as one of the potential treatments in patients with severe COVID-19 due to its ability to block IL-6-mediated inflammatory response. TCZ is an anti-IL-6 monoclonal antibody previously used in the treatment of rheumatoid arthritis and giant cell arteritis.[20] Various single-arm non-randomized studies have evaluated the effect of TCZ in severe COVID-19 and claimed a notable improvement in the clinical condition of these patients.[16, 21] **Table 1** lists currently published single-arm studies that evaluated TCZ in the treatment of severe COVID-19. Several randomized controlled trials (RCTs) are currently being conducted to evaluate the efficacy of TCZ in COVID-19.[22, 23] In this article, we performed a systematic review and meta-analysis of studies that compared the role of TCZ on mortality in severe COVID-19 as compared to the SOC.

**Table 1.**
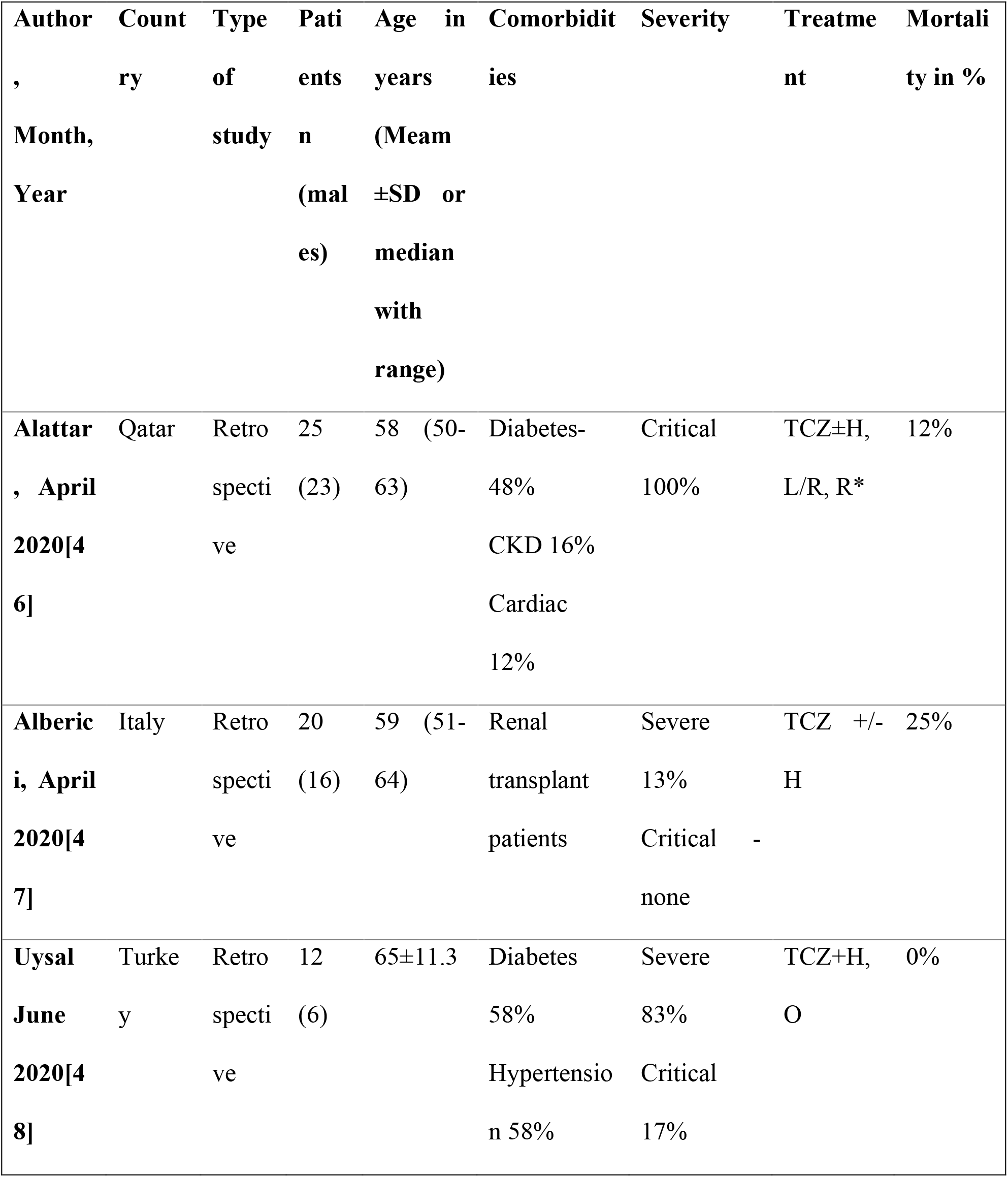
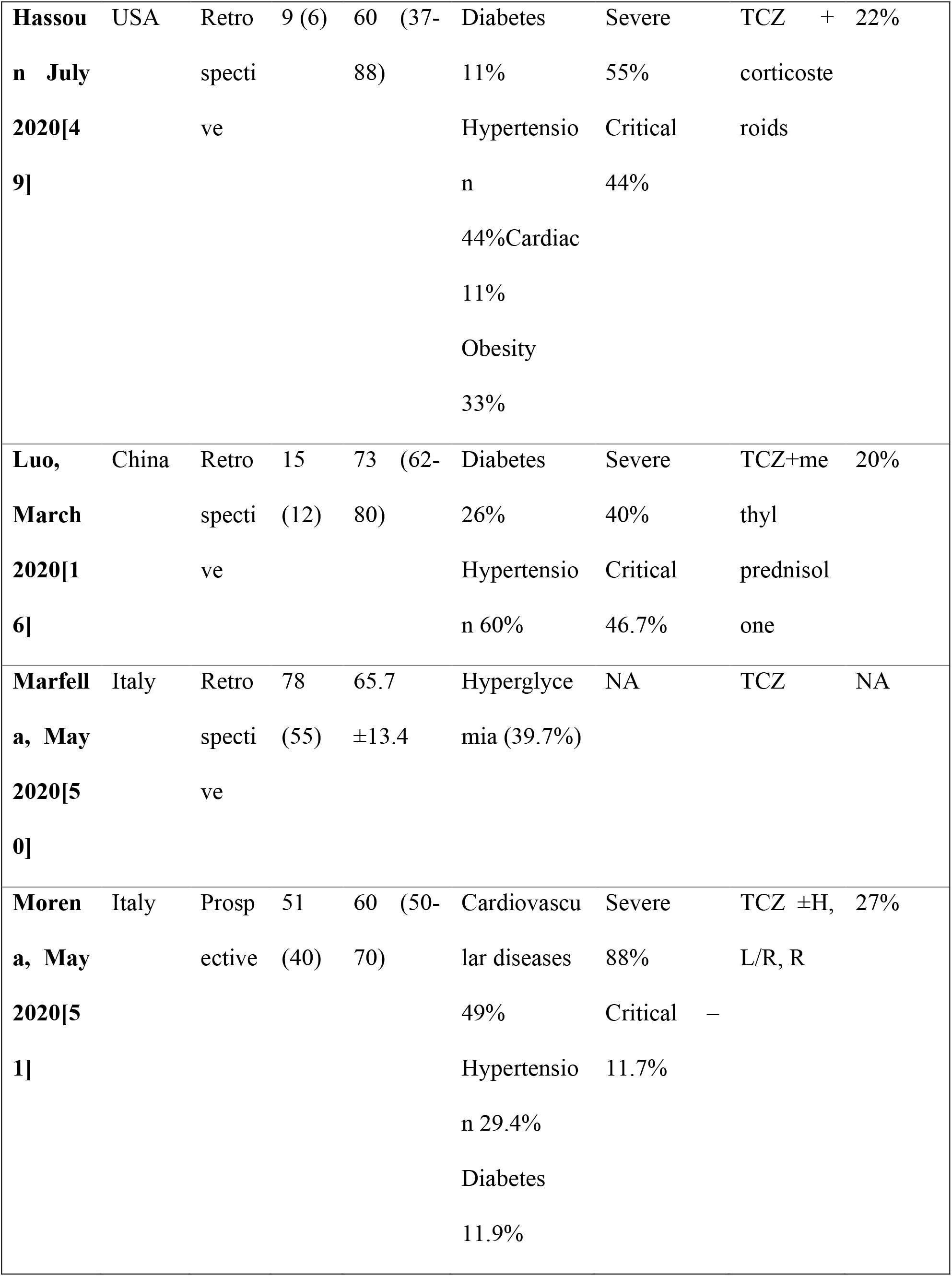
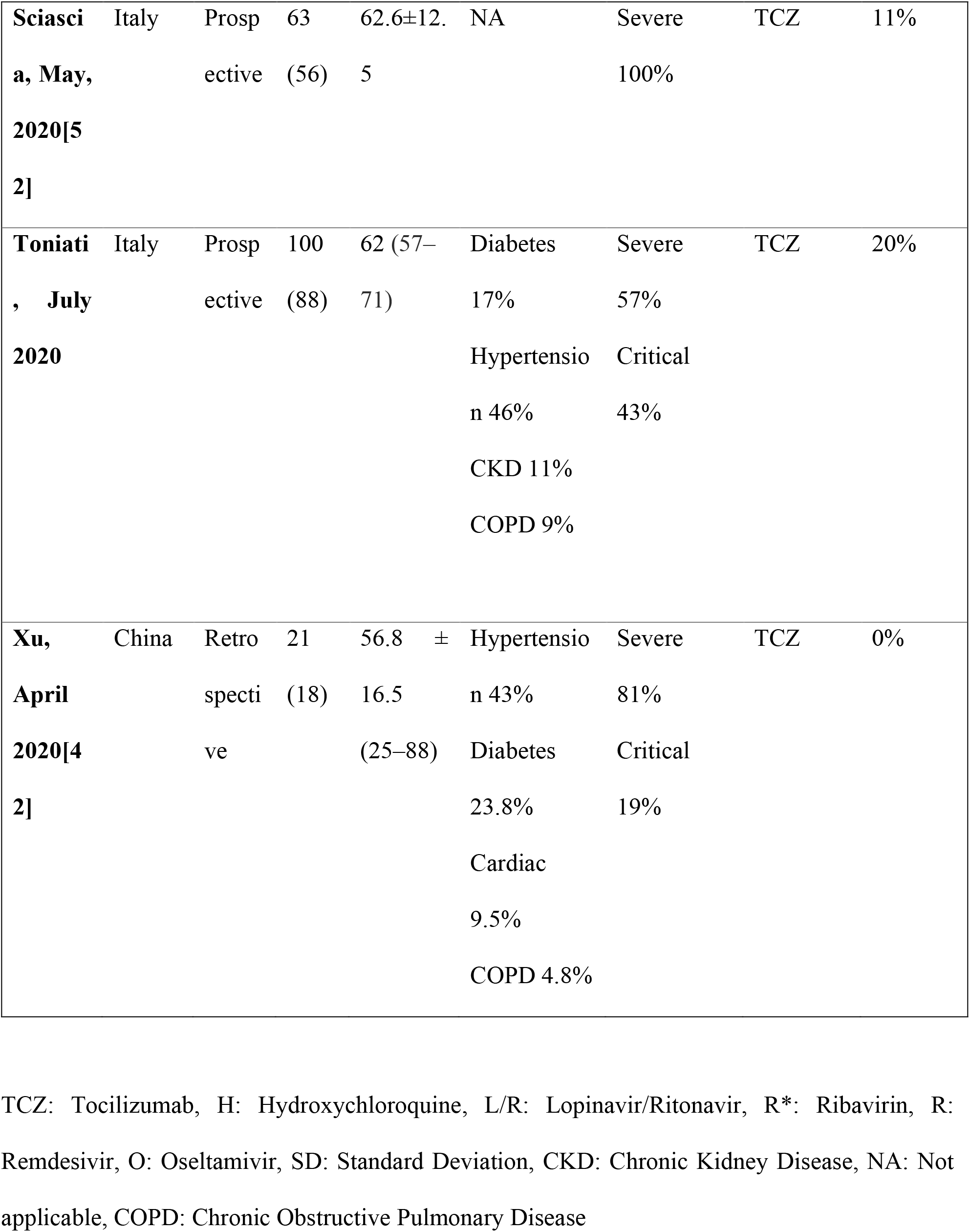
Characteristics of single-arm studies evaluating Tocilizumab in COVID 19 patients:

## MATERIALS AND METHODS

This meta-analysis was performed in accordance with Preferred reporting items for systematic review and meta-analysis statement (PRISMA).[24]

### Definitions

I. **Severe COVID-19**: Patients with respiratory rate ≥ 30 breaths/min, or peripheral capillary oxygen saturation ≤ 93% or PaO2/FiO2<=300 mmHg, or combination of these findings.
II. **Critical COVID-19**: Patients with confirmed COVID-19 who required intensive care unit (ICU) management due to mechanical ventilation or sepsis management
III. **Standard of care** (Control group): Patients with confirmed COIVD-19 were treated with any combination of the following treatment options; supplementary oxygen, antiviral agents (Remdesivir, Lopinavir/Ritonavir, Darunavir/Cobicistat), antimalarial agents (Hydroxychloroquine), anti-inflammatory agents such as steroids, and antibiotics (Azithromycin)
IV. **Tocilizumab group:** Patients were treated with TCZ in addition to SOC

**Patients, intervention, comparison, and outcomes (PICO) questions:**

**Patients:** Patients admitted to hospitals with a confirmed diagnosis of COVID-19

**Intervention:** Evaluation of TCZ in the treatment of severe COVID-19

**Comparison:** TCZ and SOC in the treatment of severe COVID-19

**Outcome:** Reduction in all-cause mortality

### Search strategy

An electronic database literature search for studies that compared ‘TCZ and SOC’ for the treatment of COVID-19 was performed on PubMed, Embase, Cochrane library, Web of Science, and MedRxiv for articles published from December 2019 till June 14^th^’ 2020. Search words ‘Tocilizumab,’ ‘anti-interleukin-6 antibody,’ and ‘COVID-19’ or ‘coronavirus 2019’ in various combinations.

### Study selection criteria

We included clinical studies that reported mortality data in severe COVID-19 patients that were treated with TCZ. The primary outcome of interest was mortality in severe COVID-19. Only studies that compared mortality rates of patients who received TCZ and SOC were included in the meta-analysis. The articles, which were in the form of only abstracts, letters without original data, case reports, reviews, meta-analysis, animal studies, or studies that did not report original data on both TCZ and SOC groups, were excluded. Further attempts were made to identify relevant studies through references to find eligible studies.

### Data extraction

Initial data collection was done by two independent investigators (UB and KN) who imported the data onto a standardized excel sheet. When there was no consensus on the eligibility of the studies, a senior author (HG) reviewed the study independently, and the final decision was made regarding inclusion or exclusion of the study. The following data were extracted from each study; authors’ names, country, type of study, number of patients in SOC and TCZ groups, patients’ gender, age, drugs used in standard of care, and mortality in both groups.

### Quality assessment

The quality of the included studies was assessed using the Cochrane risk of the bias assessment tool. Quality assessment and scoring of each included studies that were performed based on the selection of study groups, compatibility, and assessment of outcomes.

### Outcomes

The primary outcome of interest was mortality in severe COVID-19 patients in those who received SOC versus TCZ. Sensitivity analysis was performed to evaluate the effect of each study on the pooled estimates by the exclusion of one study at a time. Any significant change in pooled estimates was reported. We also performed a sensitivity analysis for studies from Europe and the USA, retrospective versus prospective studies, and studies that used antiviral medications versus those which did not. Sensitivity analysis was also done to evaluate the difference in primary endpoints when steroid was used in the SOC.

### Statistical analysis

The data extracted from studies found on online databases, including PubMed, Embase, Cochrane library, and web of science, were peer-reviewed. Studies that were retrieved from medRxiv were not peer-reviewed, and analysis performed on these studies was considered exploratory. The primary outcome of interest was pooled all-cause mortality in severe COVID-19. Categorical variables were reported in percentage and continuous variousl in mean with standard deivation. The pooled estimates with a 95% confidence interval (CI) were synthesized by meta-analysis using the ‘DerSimonian-Laird random-effects model.’ Heterogeneity across the included studies was performed using inconsistency index I^2^. The heterogeneity was classified as low, moderate, and substantial heterogeneity, when the inconsistency index was 25%, 50%, and 75%, respectively. The publication bias was assessed using funnel plot analysis and Egger’s test. Statistical analysis was done using Review Manager (Version 5.3; The Nordic Cochrane Centre, Copenhagen, Denmark) and STATA 14.2 (StataCorp. 4905 Lakeway Drive, College Station, Texas 77845 USA).

## RESULTS

### Study characteristics

A total of 693 studies were identified through an initial literature search. After exclusion of duplicates and screening of articles based on the title and abstract, 51 articles remained. These articles were further reviewed thoroughly, and finally, sixteen studies were eligible for the primary outcome. There were thirteen retrospective studies[17, 25-36] and three prospective studies.[37-39] **Supplementary Figure 1** provides details of the literature search and study selection process.

There were a total of 3,641 patients with 64% males from sixteen included studies. There were 2,488 patients (61.7% males) in the SOC group and 1,153 patients (68.7%) in the TCZ group. All patients included in the meta-analysis received SOC, and the TCZ group received Tocilizumab in addition to SOC. Hydroxychloroquine was used in all the included studies, Azithromycin was used in 6 studies, Lopinavir/Ritonavir combination was used in 6 studies, steroids were used in 12 studies, Darunavir and Cobicistat combination was used in 3 studies, and remdesivir was used in 2 studies. **Table 2** lists the studies included in the meta-analysis and study characteristics.

**Table 2:**
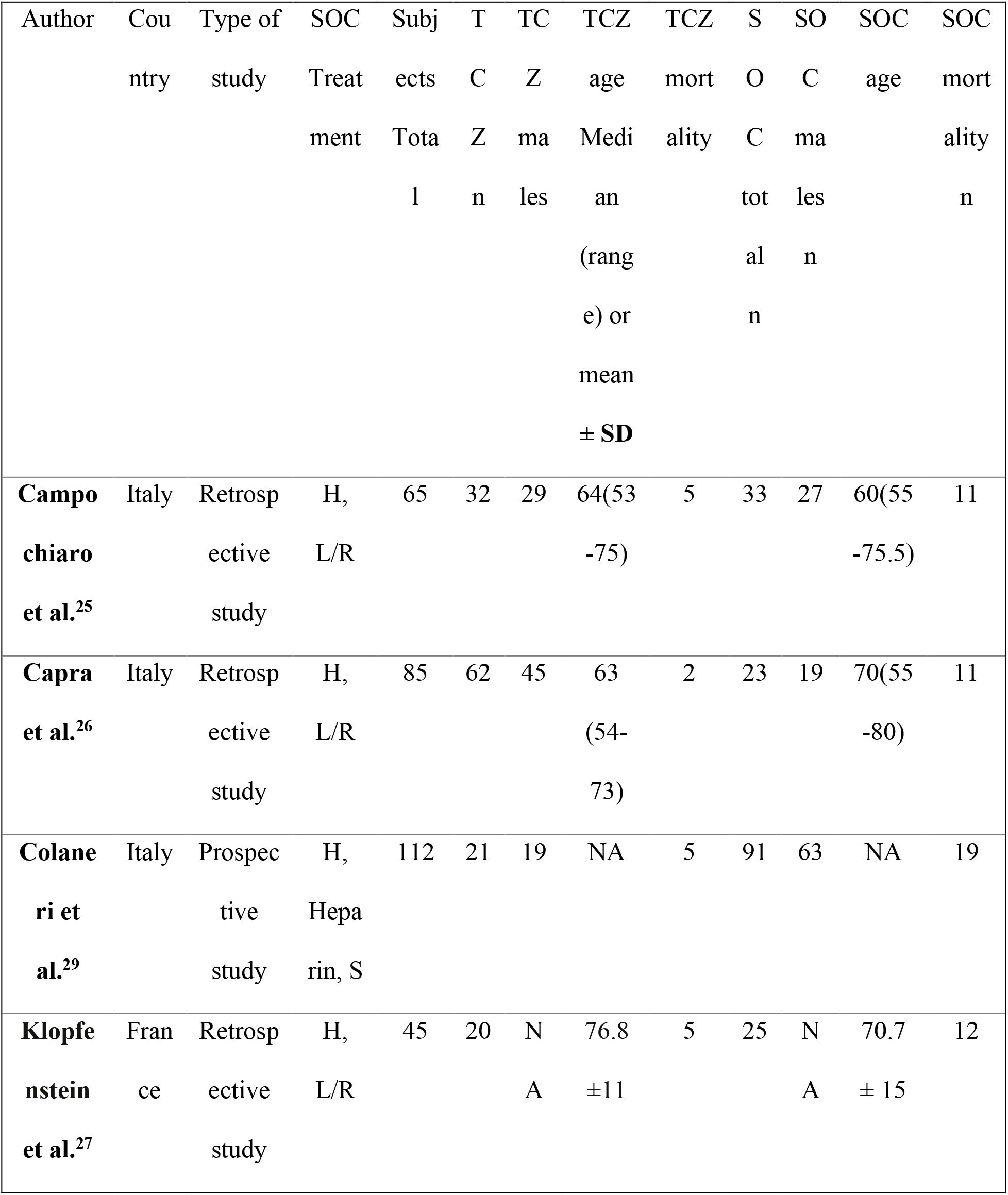
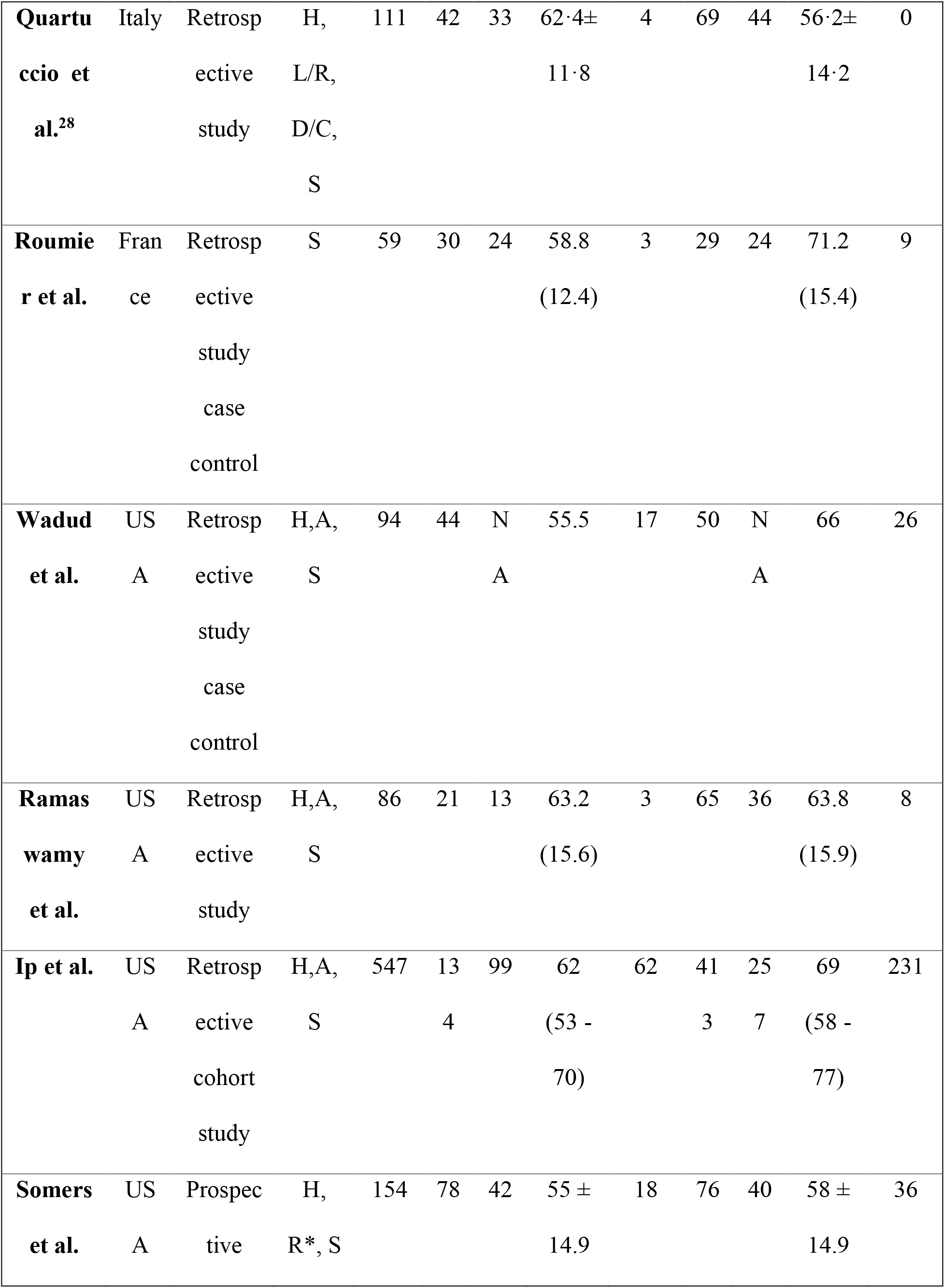
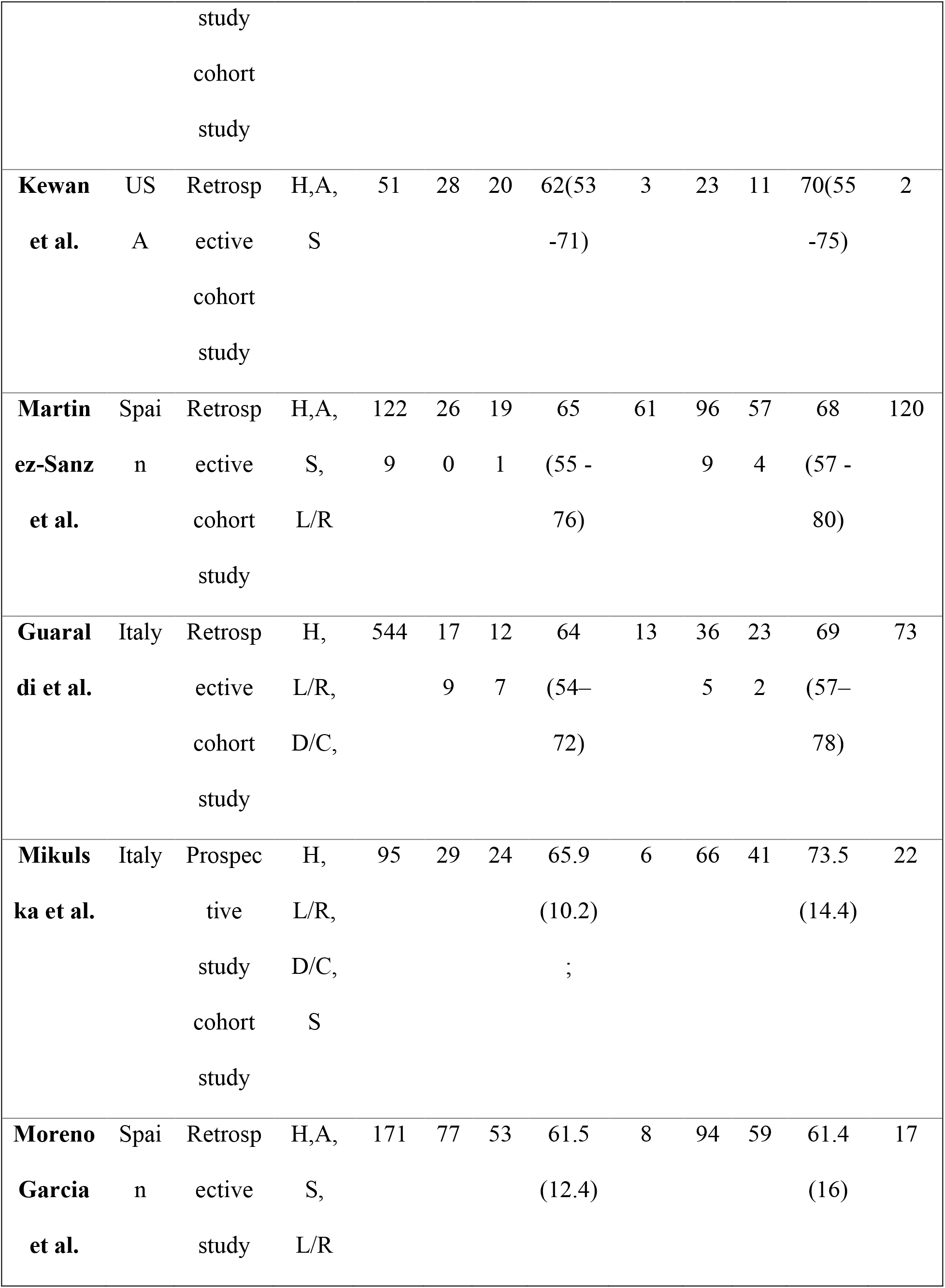
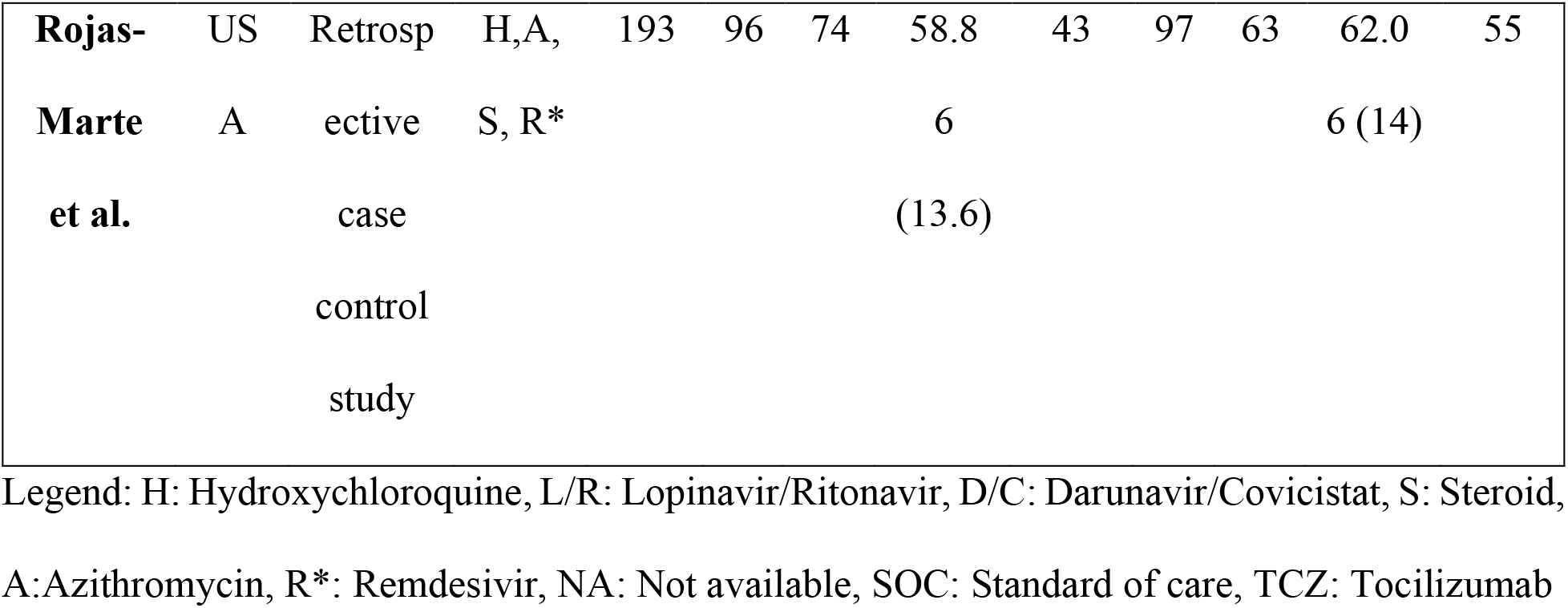
Characteristics of studies included in the meta-analysis

### Primary outcome and sensitivity analysis

The mortality rate of COVID-19 patients in the TCZ group was 22.4% (258/1,153), and the mortality rate in the SOC group was 26.21% (652/2488). The pooled odds ratio was 0.57 (95% CI 0.36-0.92; p=0.02). Forest plot analysis of the primary outcome is shown in **Figure 1**. There was substantial heterogeneity among the included studies (I^2^=80%).

**Figure 1:**
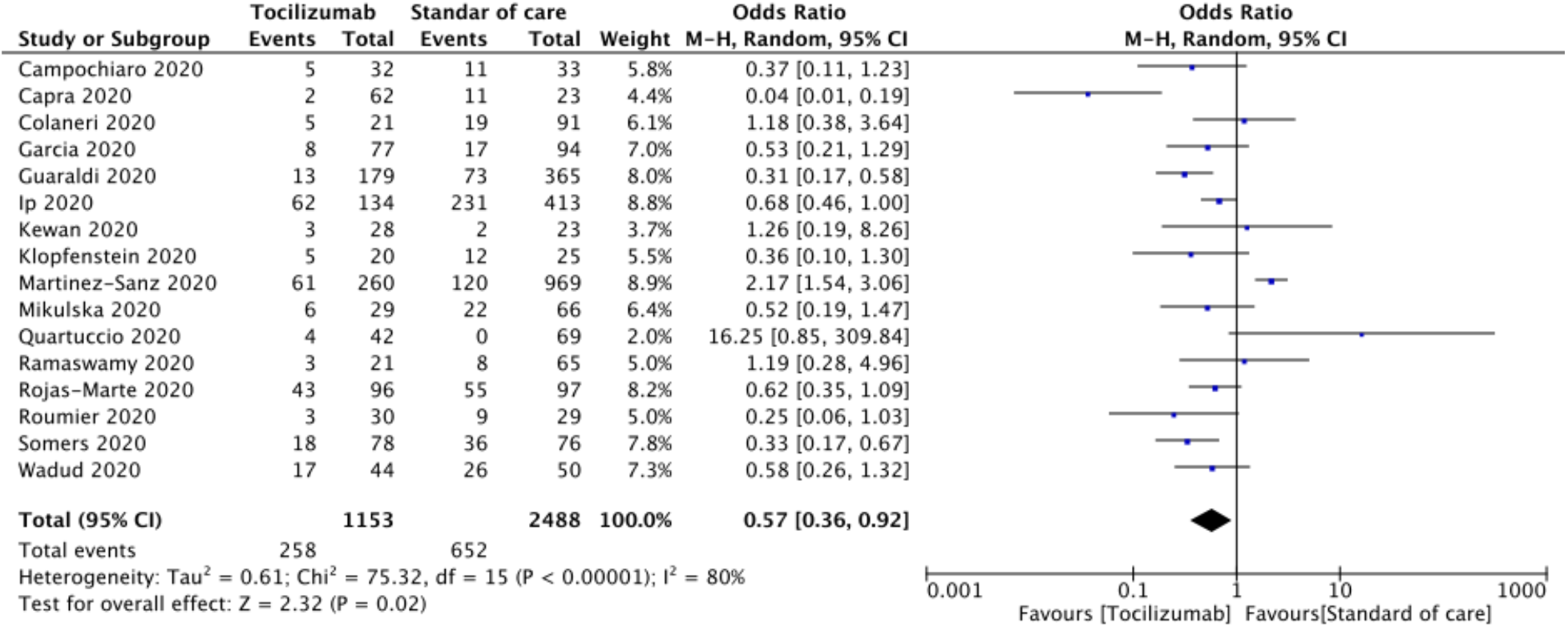
Forest plot comparing patients treated with Tocilizumab versus Standard of care alone

Sensitivity analysis by the exclusion of each study at a time showed no significant difference except when Capra et al.[26] excluded; loss of statistical significance was observed (Pooled OR 0.65 [95% CI 0.42-1.01] p=0.060). Sensitivity analysis of retrospective and prospective studies showed pooled OR 0.57 (95%CI 0.33-0.99; p=0.05), and pooled OR 0.53 (95% CI 0.26-01.09; p=0.08), respectively. **Figure 2** shows forest plot analysis for the pooled estimations of retrospective and prospective studies. Sensitivity analysis of studies that used steroid as SOC and those did not show a pooled OR 0.76 [95% CI 0.47, 1.23; p=0.27] and pooled OR 0.24 [95% CI; 0.10-0.54, p<0.01], respectively. **Figure 3** shows the forest plot analysis of sensitivity analysis for the use of steroids in the included studies. Further, sensitivity analysis of studies from Europe and USA showed pooled OR 0.52 [0.23, 1.17; p=0.12] and pooled OR 0.61 [0.46, 0.79; p<0.01], respectively. Supplementary figure 2 shows forest plot for sensitivity analysis of the American and

**Figure 2:**
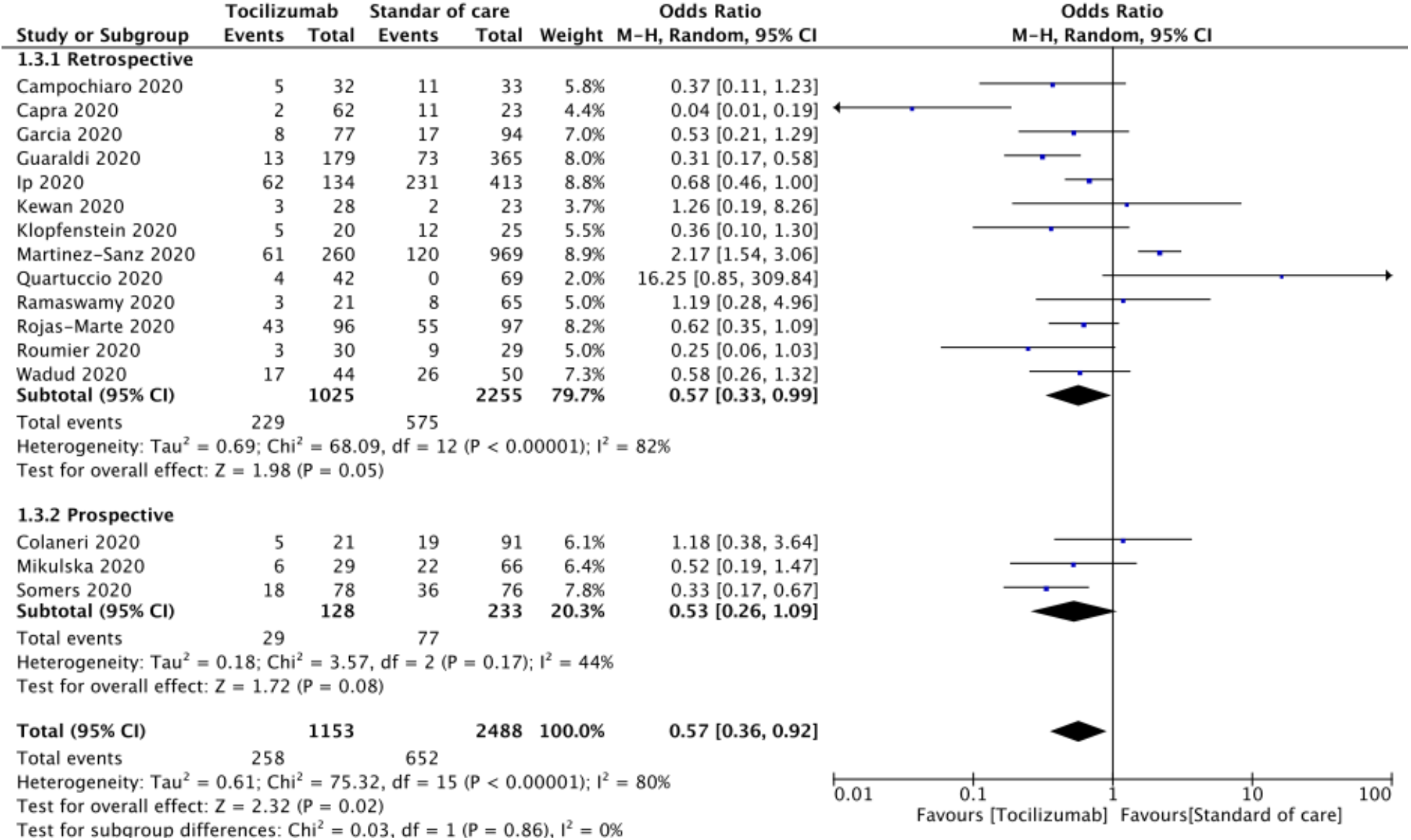
Forest plot for sensitivity analysis of retrospective and prospective studies

**Figure 3:**
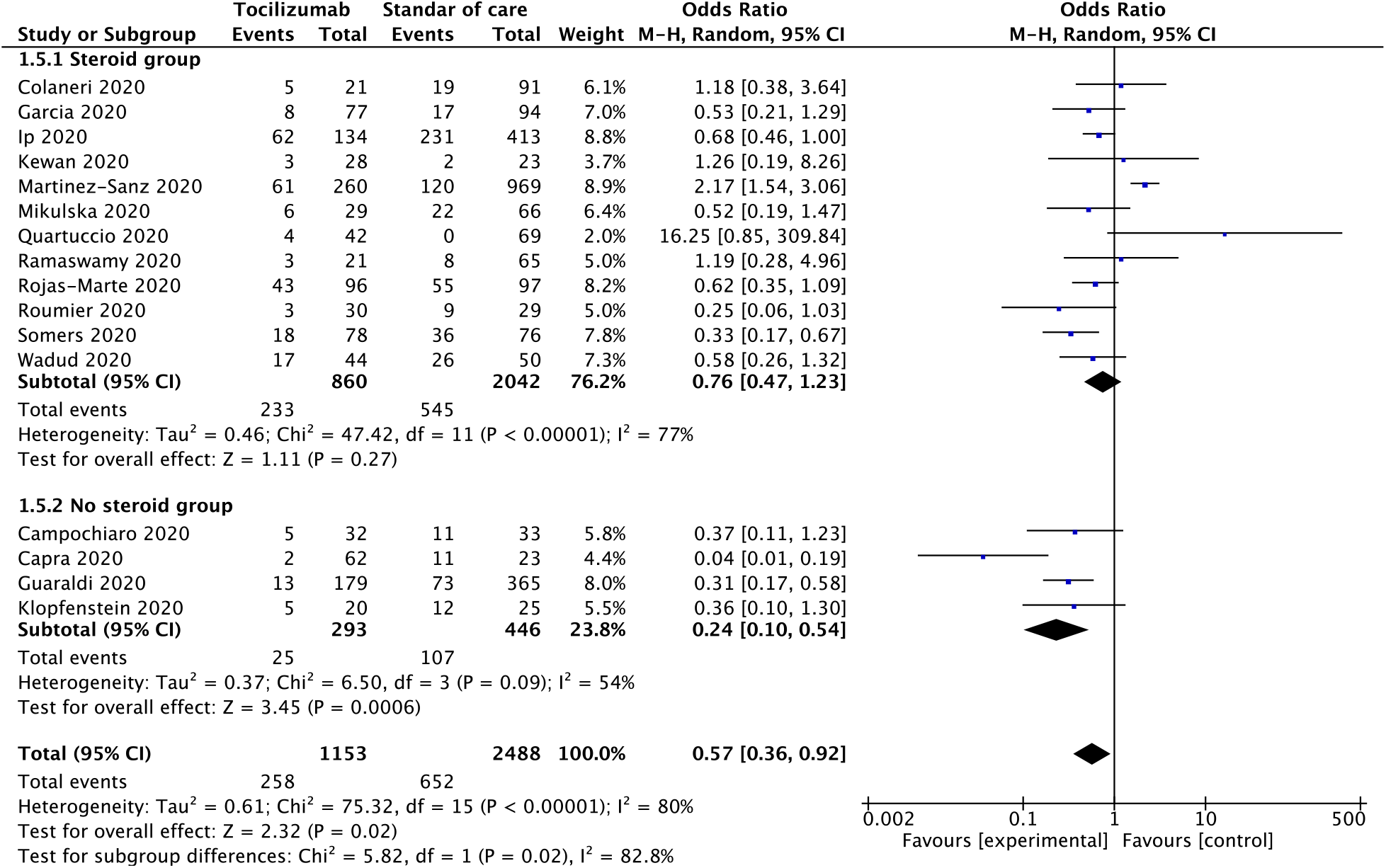
Forest plot for sensitivity analysis of steroid use in SOC (Random-effects model)

European studies. Quality assessment of included studies was done using the Cochrane risk of bias tool with parameters for each study is shown in **Supplementary Table 1**.

### Publication bias

Analysis of publication bias using the funnel plot showed the clustering of studies towards the peak and median (**Supplementary Figure 3**). There were four studies that were outside of the funnel. Further analysis using Egger’s test (p = 0.087) and Begg’s test (Z =0.09) suggested no significant publication bias.

## DISCUSSION

In this meta-analysis including 3,641 patients, TCZ (11.9%) group had 3.81% less number of deaths when compared to SOC (n=16; pooled OR 0.57 [95% CI 0.36-0.92] p=0.02). The analysis also showed that, when steroids are used in the treatment of severe COVID-19, there was no statistical difference in mortality between TCZ and SOC (n=12; pooled OR 0.76 [95% CI 0.47, 1.23; p=0.27). However, when steroid was not used TCZ group had significantly low mortality when compared to the SOC group (n=4; pooled OR 0.24 [95% CI; 0.10-0.54] p<0.01). Sensitivity analysis showed that mortality trends in prospective studies suggest TCZ group did not show a significant difference when compared to SOC (n=3; pooled OR 0.53 (95% CI 0.26-01.09; p=0.08). This is likely due to a lack of power since there were only three studies with the small study population. Possible reasons for substantial heterogeneity among the studies included in the meta-analysis (I^2^=80%) are; the difference in the age and comorbidities, variability in the follow-up period, and definition of death due to COVID-19, and response to treatment.

Our meta-analysis shows that the addition of TCZ might significantly reduce mortality in severe COVID-19. However, when steroids are used in the standard of care, the absence of a significant difference in mortality suggests possible role anti-inflammatory properties of steroids in the treatment of severe COVID-19. It was also observed that studies from Europe showed no significant difference in mortality between the TCZ group and the SOC group (n=10; pooled OR 0.52 [95% CI 0.23-1.17] p=0.12). However, Studies from the USA did show lower mortality in the TCZ group when compared to SOC (n=6; pooled OR 0.61 [95% CI 0.46-0.79] p<0.01). This could be related to an acute shortage of ICU beds in Italy and a rapid rise in COVID-19 cases in Italy since most of the European studies were from Italy. This also highlights the availability of resources can significantly affect the outcomes.

SARS-CoV-2, the causative organism of COVID-19, commonly enters the human body through inhalation of droplets. Once the virus comes in contact with respiratory mucosa, the spike (S) protein on the viral cell membrane fuses with the angiotensin-converting enzymes 2 (ACE2) receptors expressed on the mucosal cells, and the viral RNA enters the cell.[40] These infected cells undergo apoptotic changes and phagocytosed by macrophages and presented to antigen-presenting T cells, which in turn leads to CD4 T cell-dependent immune response and results in increased antibody production from B lymphocytes. Ultimately, the unrestricted release of inflammatory cytokines from the immune response leads to ‘cytokine release syndrome’ (CRS). Patients with severe COVID-19 have been shown to have lymphocytopenia and increased plasma concentration of pro-inflammatory cytokines such as IL6, interleukin 10, granulocyte stimulating factor (G-CSF), monocyte chemoattractant protein 1 (MCP-1), tumor necrosis factor (TNF).[41]

The role of TCZ in the treatment of COVID-19 is hypothesized based on its ability to block IL 6 activity, a pro-inflammatory cytokine that plays a significant role in the development of ARDS. In a prospective single-arm observational study by Xu et al., twenty-one patients were treated with TCZ. At the end of 5 days, there was a significant improvement in the inflammatory level of markers, lymphocytopenia, and interstitial lung changes—oxygen requirement reduced in all of the patients. None of the patients died.[42] In another prospective single-arm study by Toniati et al., 100 patients with critically ill COVID-19 patients requiring mechanical ventilation were treated with TCZ. At the end of 10 days follow up, 77% of patients experienced significant improvement in respiratory condition and clearing of bilateral interstitial opacities on chest X-ray. The study showed overall mortality of 20%. In another single-arm observational study, 63 hospitalized patients with COVID-19 treated with TCZ also had improvement in oxygen requirement, with an increased likelihood of survival (Hazard ratio 2.2; 95%CI 1.3, 6.7; p<0.05) and 11% overall mortality. Several other studies showed that TCZ might have a beneficial effect in the treatment of COVID-19 in organ transplant recipients.[43-45] Based on these encouraging results, several RCTs are underway to evaluate the efficacy of TCZ in COVID-19.

Although our study shows that there is no clear evidence to show the superiority of TCZ over SOC, nearly 45% less death in the TCZ group suggests possible benefit in reducing the mortality in severe COVID 19. Future studies should consider evaluating TCZ against SOC and convalescent serum in the management of severe COVID 19. It is also important to note that TCZ may have a supporting role in combination with SOC, and this needs to be explored as well.

Our study has a few limitations that need to be noted; most of the included studies are retrospective observational studies with only three prospective studies. Most of the included studies were from Italy and the United States. Therefore, the results may not be generalizable. The risk of selection bias is high since most studies did not report whether patients included in the study were consecutive. We were unable to estimate the effect of resource shortage on the mortality, since there was a reported shortage of intensive care units to provide appropriate care for severe COVID-19 patients in Italy. Some of the severe COVID-19 patients were treated in the general ward. Mortality can be significantly affected by comorbidities, which is not evaluated in our meta-analysis. This may have played a role in patient mortality and, therefore, might have affected the outcomes. It should be noted that the administration of antiviral agents was not uniform, and not all pateints received the same antiviral agents in either TCZ group or SOC group. Therefore, the findings of this meta-analysis needs to be validated in well designed randomized clinical trials. Also, we were unable to determine which patients would benefit from TCZ and which patients do not. Lastly, pooled analysis of studies with substantial heterogeneity is debatable. Large RCTs with sufficient power to evaluate the effectiveness of TCZ in the treatment of COVID-19 are needed. Also, there appears to be a role for steroids in the treatment of severe COVID-19. Future studies should evaluate TCZ versus steroids in treating severe COVID patients in randomized clinical trials.

In conclusion, this systematic review and meta-analysis summarize that the addition of TCZ to SOC might reduce the mortality in severe COVID-19. With no definitive treatment available, TCZ might be a choice in the treatment of severe COVID-19. Large, high-quality prospective studies and RCTs are urgently needed to evaluate the role of TCZ in the treatment of severe COVID-19.

## Data Availability

The data referred to in this manuscript is available freely on online databases mentioned in the manuscript

## Acknowledgements

None

## Declarations of interest

None

## Contributions

Conception and design: Umesha Boregowda, Hemant Goyal

Literature review: All authors

Drafting manuscript: Umesha Boregowda, Kamala Nanjappa, Mahesh Gajendran

Critical revision and editing: All authors

Final approval: All authors

## Funding

There was no funding for this research

**Abbreviations**
COVID 19: coronavirus disease 2019
SARS: Severe Acute Respiratory Syndrome
SARS-CoV-2: Severe acute respiratory syndrome coronavirus-2
ARDS: Acute respiratory distress syndrome
OR: Odds ratio
I^2^: Inconsistency index
TCZ: Tocilizumab
SOC: Standard of care
RCT: Randomized controlled trials
G-CSF: Granulocyte stimulating factor
MCP-1: Monocyte chemoattractant protein 1,
TNF: Tumor necrosis factor
ACE2: Angiotensin-converting enzymes 2
CRS: Cytokine release syndrome
SD: Standard Deviation
CKD: Chronic Kidney Disease
NA: Not applicable
COPD: Chronic Obstructive Pulmonary Disease
TCZ: Tocilizumab
H: Hydroxychloroquine
L/R: Lopinavir/Ritonavir
R*: Ribavirin
R: Remdesivir
O: Oseltamivir
S: Spike protein

---

**Supplementary Figure 1:**
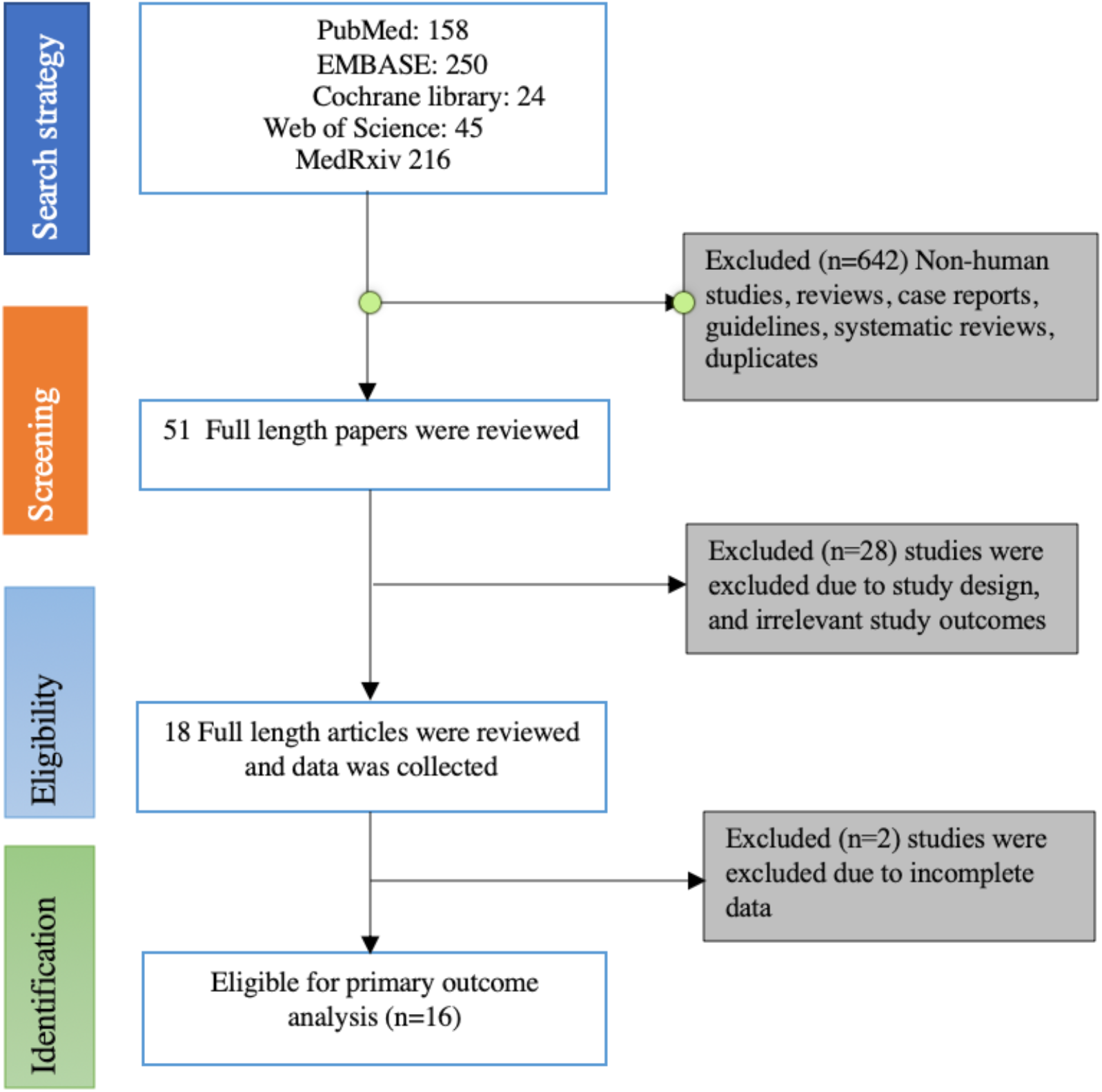
Flow chart of selecting eligible studies for the meta-analysis

**Supplementary Figure 2:**
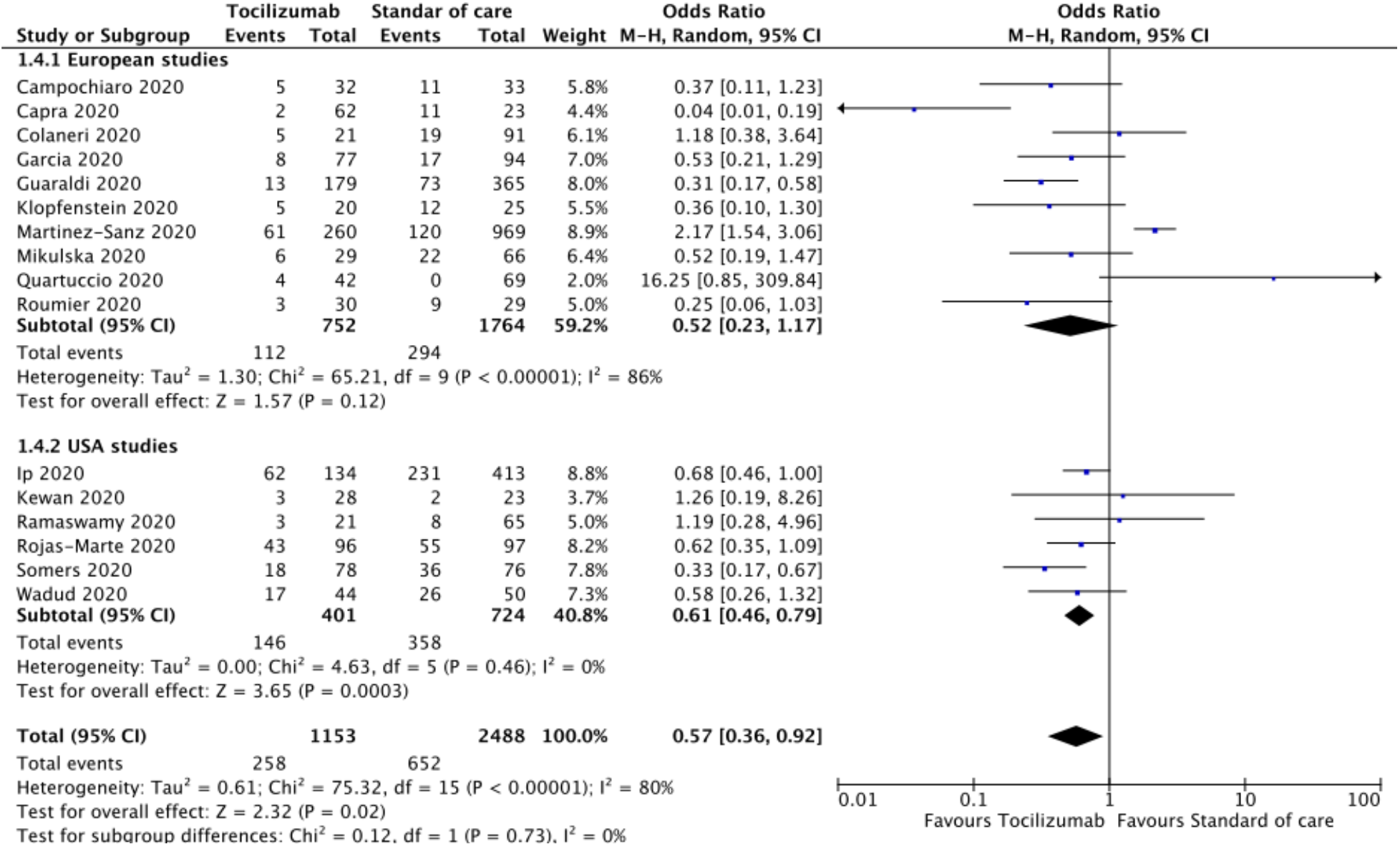
Forest plot for sensitivity analysis for the American and European studies

**Supplementary Figure 3:**
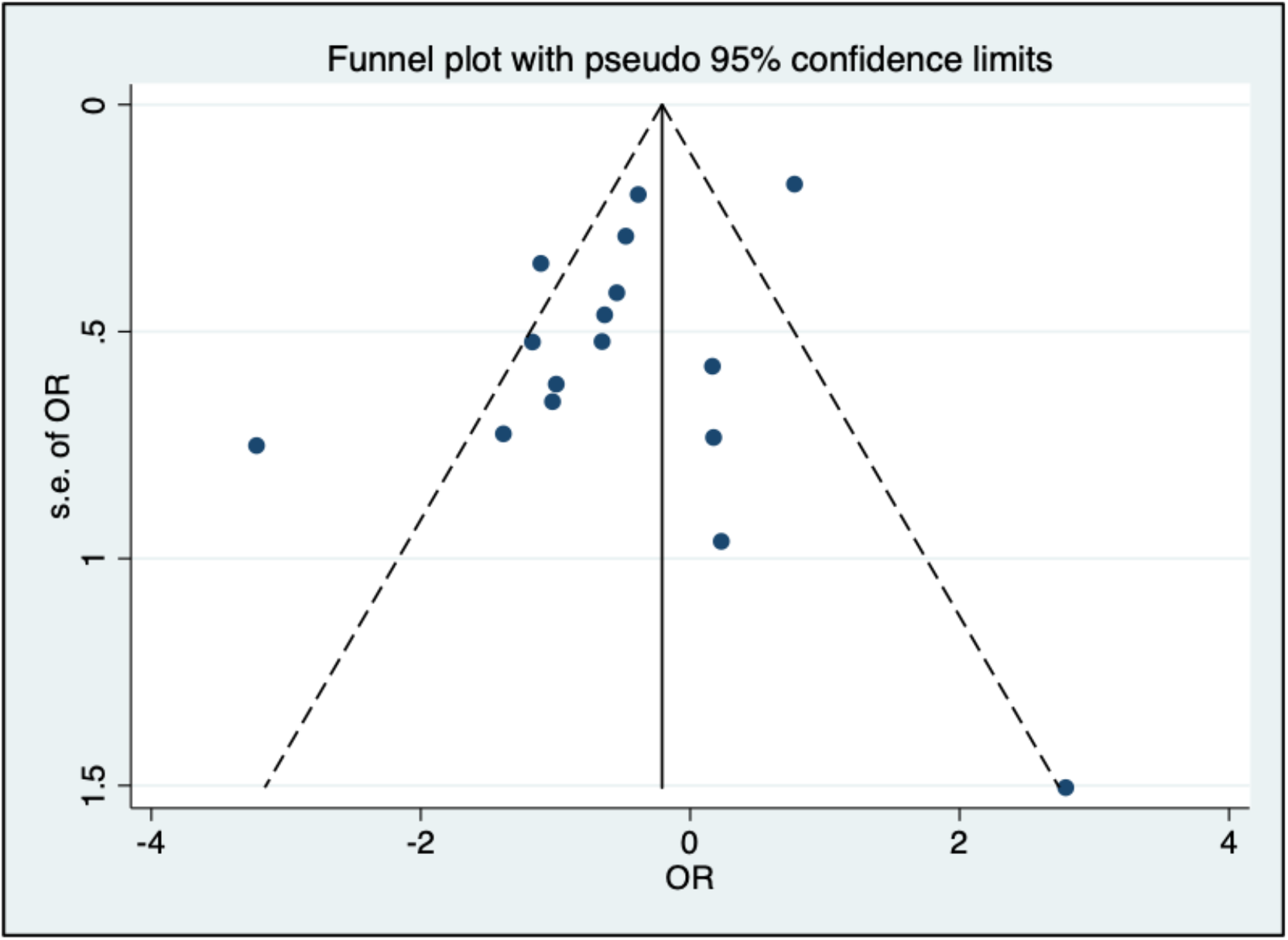
Funnel plot analysis of publication bias

**Supplementary Table 1:**
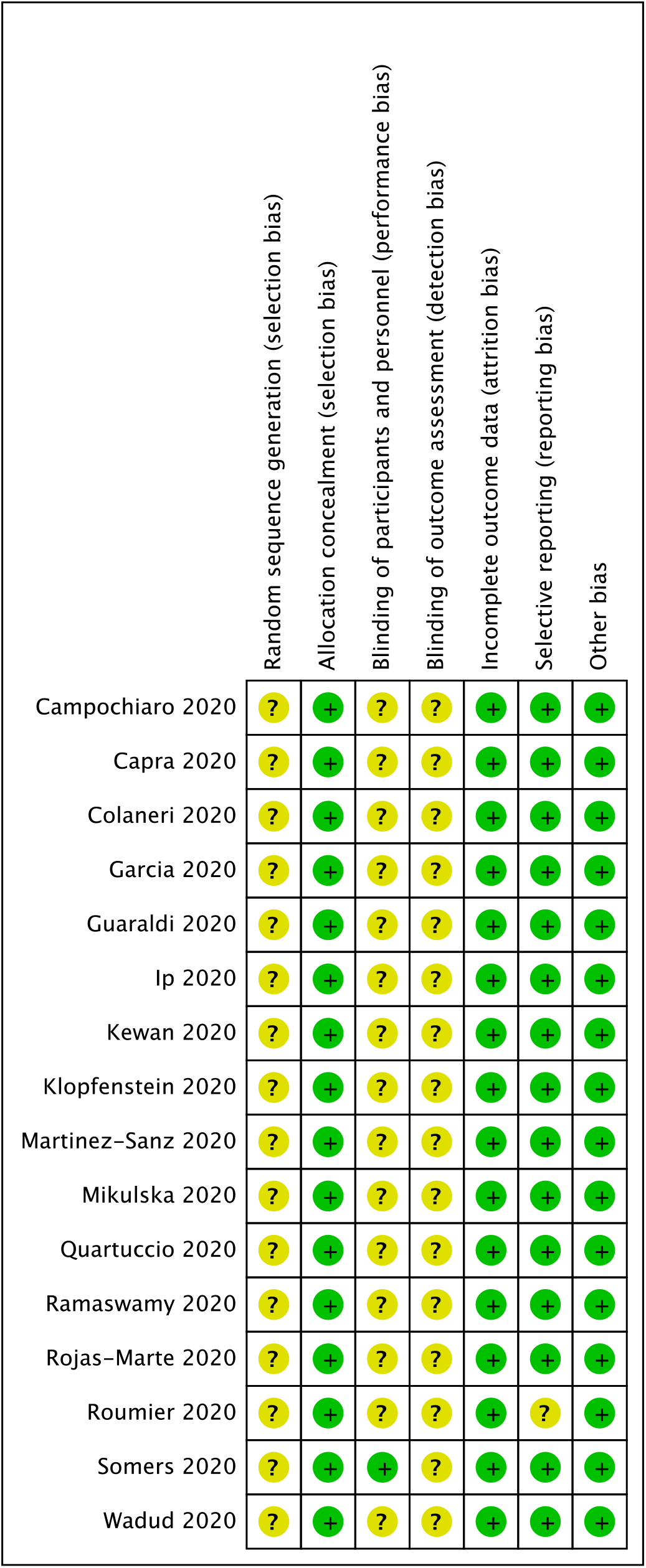
Cochrane risk of bias table

## Notes

### Competing Interest Statement

The authors have declared no competing interest.

### Funding Statement

There was no funding available for this study

### Author Declarations

This research did not require IRB approval since it is a literature review

### Summary of Updates

Results section of the abstract was updated to reflect the correct number of studies included in the meta-analysis.

